# Evaluating Genetic-Based Disease Prediction Approaches Through Simulation

**DOI:** 10.1101/2025.03.21.25324431

**Authors:** Max Shpak, Eric Parfitt, Soroush Mahmoudiandehkordi, Mehdi Maadooliat, Steven J. Schrodi

## Abstract

Common diseases exhibit substantial heritability, and GWAS of these diseases have revealed hundreds of thousands of high-frequency disease susceptibility variants throughout the genome. These studies offer the prospect of using genomic data to improve disease prediction and diagnosis, however, the relative performance of different predictive modeling approaches is not well-characterized. To investigate this systematically, we constructed a Monte Carlo simulation generating model genomes with large numbers of SNPs, with a proportion of SNPs carrying risk alleles that are parameterized by the strength of their effects and by different modes of inheritance – additive, dominant, recessive, and combinations thereof. After generating genotypes for cases and controls, several machine learning classifiers (logistic regression, naïve Bayes, random forests, and neural networks, with and without feature selection) were applied to predict disease phenotype from genotypes. Each classifier’s rates of false positives and false negatives were evaluated and compared using AUC. We found that random forest models were the most accurate predictors of disease phenotype over the range of inheritance parameters, followed by logistic regression and naïve Bayes, while the feedforward multilayer neural network-based predictive model had lower AUC. Furthermore, with the small fraction of null sites in our model, there was almost no difference in the performance of classifiers with or without LASSO-based feature selection. We also investigate the association of AUC with the difference in polygenic risk score (PRS) between disease and control samples by comparing AUC in the simulations to the values predicted from the PRS distributions based on odds-risk and liability models.

## INTRODUCTION

Large-scale genetic association studies of complex diseases have markedly improved our understanding of the specific variants, genes, and pathways underlying a wide array of disease phenotypes and medically relevant traits (Beck et al 2020, Visscher et al 2018). These discoveries have played an instrumental role in 1) improving our understanding of inheritance patterns and genetic architecture of complex diseases (Watanabe et al 2019), 2) providing insight into the roles of protein function, splicing, and gene regulation in disease (Maurano et al 2012, Gallagher and Chen-Plotkin 2018), 3) identifying genetic signatures of therapeutic response (McInnes et al 2021), 4) identifying drug targets and mechanisms (El-Husseini et al 2020, Levey et al 2021, Owen et al 2010, Reay and Cairns 2021, Tachmazidou et al 2019), and 5) have illuminated paths to better predict disease risk (Schrodi et al 2014).

The use of genetic factors in disease prognosis is one of the key promises of modern human genomics. Identifying risk-predictive combinations of germline variants not only better assesses the likelihood that patients will develop a disease, it can also shift clinical diagnoses to earlier stages in the healthcare system, thereby providing opportunities for intervention prior to excessive morbidity. Early clinical applications of genetic information include neonatal screening programs (Baker et al 2019, Farnaes et al 2018), hereditary cancers (Burt and Neklason 2005, Samadder et al 2020), and pharmacogenetics (Relling et al 2019, Roden et al 2018). Many of these initial efforts were understandably focused on monogenic traits with variants carrying very high effect sizes.

Researchers have also developed and implemented methods to combine signal from panels of disease susceptibility markers to predict complex diseases. One of the first statistically rigorous approaches to using an arbitrary number of disease-associated genetic markers for the purpose of prediction of disease traits was outlined by Yang et al 2003. The approach used in their study was to calculate the likelihood ratio of the probability of a panel of multilocus genotypes in case (disease) vs. control (healthy) individuals. The likelihood ratio was estimated by a logistic regression model, enabling adjustment for covariates and interaction effects, which can then be used to calculate the positive and negative predictive values of disease using multilocus genotypes. Along similar lines, a number of disease risk metrics have been developed based on genotype effects—among the most widely adopted approaches is the construction of polygenic risk scores (PRS), which are a weighted average of the per-SNP log-transformed odds ratios taken over a number of sites (Dudbridge 2013, Janssens et al 2006, Purcell et al 2009, Schizophrenia Working Group 2014).

Machine learning and deep learning algorithms are robust, powerful, and established methods for addressing classification problems, and offer alternatives to PRS-based methods of assessing disease risk from genotype data. Recently, machine learning tools have been applied to genetic data for the purpose of disease prediction (Ho et al 2019, Telenti et al 2018, Wu et al 2018). These methods include classic machine learning classifiers, such as random forests and support vector machines, as well as a variety of newer neural network-based algorithms such as convolutional neural networks, generative adversarial networks, and self-organizing maps (e.g. Alzoubi et al 2023, Bracher-Smith et al 2021, Ghafouri-Fard et al 2020, Patrick et al 2018).

As genomic technologies advance and large-scale disease gene mapping studies continue to proliferate, the resulting wealth of genetic association data offers an opportunity to construct genetic-based and multi-omics-based predictive models for disease traits. However, little is known about the performance of these predictive models under different genetic architectures, or how these performances compare to other genotype-based disease risk assessment metrics. Allele frequencies, genotype penetrances, the number of susceptibility loci, and epistatic interactions all play a role in the genetic architecture of diseases. Understanding the impact of these parameters on the performance of various types of predictive modeling efforts is critically important for improving the accuracy of disease prediction from genomic profiles. For example, early work showed the importance of genotype frequency on discrimination accuracy in the context of type 2 diabetes (Janssens et al 2007) and additional work examined the comparison between multiplicative and additive modes of inheritance (Moonesinghe et al 2011).

It is plausible that different prediction algorithms have different strengths and weaknesses in their ability to capture disease susceptibility effects from combinations of SNPs, and that their relative predictive accuracies depend on the genetic architecture underlying disease risk. Understanding how the performance of various predictive methods changes as a function of disease genetic parameters will provide insight into which class of models will have the highest predictive accuracy for specific medical traits with disparate genetic architectures. The goal of this work is to explore the diagnostic utility of commonly used binary classifiers as a function of parameters for modeling disease genetics. With this aim, we developed a general disease multilocus genetic model which is implemented as a Monte Carlo simulation where we generate combinations of disease susceptibility SNPs/SNVs together with sites that are null with respect to disease and assess the efficacy of various classifier algorithms at predicting disease risk from genotypes.

Because patient risk is often evaluated using heuristic metrics, it is also important to assess the extent to which such metrics correlate with the accuracy of prediction models. As noted above, PRS have become a widely used approach to combine disease association signals from multiple susceptibility markers (Choi et al 2020, see also De La Vega and Bustamente 2018, Janssens 2019, Lewis and Vassos 2020 for discussions of applicability and limitations). PRS can also be used as a summary score to concurrently test for phenotype association against the combined effects of multiple SNPs. However, the efficacy of PRS as a predictor of disease incidence from genotype has not been fully assessed through simulation, particularly not in the context of how differences in PRS between disease and control groups scale with the sensitivity and specificity of disease prediction under different classifier algorithms. Among the goals of this study are to assess the relationship between PRS values and predictive accuracy, and to use simulations to test analytical models of this relationship.

## MATERIALS AND METHODS

### Disease Model

The disease model employed simulated SNPs which varied in allele frequency and in the penetrances of their risk effects. This allows for modeling of both disease susceptibility SNPs and SNPs that are null with respect to disease status. Denoting alleles segregating at a modeled biallelic SNP as *A*_1_ and *A*_2_ and defining the genotype penetrances as *f_ij_* = *P*(*Disease*|*A_i_A_j_*), the frequency of the genotypes in the disease and non-disease populations is then

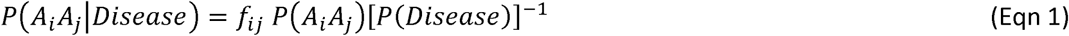

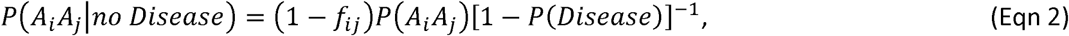

respectively. The probability of disease will be the proportion of the disease trait attributable to the specific SNP, and calculated by *∑f_ij_P*(*A_i_A_j_*).Hardy-Weinberg equilibrium is assumed in the general population.

For the purposes of this study, we consider a simplified genetic architecture and ignore epistatic interactions, so that the contribution of each site to disease risk is statistically independent of other sites. Therefore, the probability of a genotype conditional on disease (case) or control status at one locus is also independent of the genotype conditioned on disease or control at all other sites in the genome. The mode of inheritance is defined through simple relationships between penetrances of risk allele effects. Three standard models of inheritance were examined in the simulation: recessive, dominant, and additive. Although the penetrance relationships can depart from these simple models in complex diseases due to epistatic effects, evaluating these models serves as a step to explore the relative diagnostic utility across several types of genetic architecture. The recessive, dominant, and additive models are defined in terms of the penetrance relationships:

Recessive: *f*_11_ = *f*_l2_, *f*_22_ = γ*f*_l2_

Dominant: *f*_22_ = *f*_12_ = γ*f*_12_

Additive: *f*_22_ = (2γ −1)*f*_11_, *f*_l2_ = γ,*f*_11_,

where γ > 1 is the genotypic relative risk (we do not simulate the case of γ < 1, i.e. “protective” sites with alleles that reduce disease risk relative to a wild-type baseline in this study). In the additive model, the heterozygote is, by definition, exactly intermediate in disease risk between wild-type baseline and the genotype homozygous for risk alleles.

This general disease genetics framework has been used in numerous previous studies (Nielsen et al. 1998, Sham 1998, Schrodi et al 2007, Maadooliat et al 2016). We will also consider genetic architectures where the model genomes are composed of a mixture of effects across sites, i.e. the majority of sites with additive effects, and a smaller subset of sites with recessive and/or dominant effects in order to assess whether such mixed models were determined by the minority fraction of sites with dominant (or recessive) effects, versus being intermediate in comparison to additive and non-additive scenarios. Null genetic markers are modeled by setting all risk penetrances to a single constant value *f*, versus risk markers with variable perturbation term r. All SNPs were generated in linkage equilibrium to model the effects from susceptibility and null loci randomly distributed across the genome. Both large-scale genetic studies and population genetics theory have shown that the site frequency spectra is accurately modeled by Beta densities, hence, the allele frequency at each site is sampled as a Beta variate (Ewens 1979, Gudmundsson et al 2021). The two shape parameters for the Beta density (â = 1.9195, 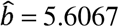 corresponding to an allele frequency expectation of 0.255 and variance = 0.0223) were estimated using the method of moments applied to all statistically significant SNPs from the GWAS Catalog (Buniello et al 2019). Allele frequencies are assigned independently across sites in our model, in keeping with the assumption of approximate linkage equilibrium between pairs of sites.

### Polygenic Risk Score Calculation

Polygenic Risk Score (PRS) calculation is an effective approach to capturing disease association across multiple genetic markers using a single univariate metric. A PRS is a weighted average of the effect size across genetic markers (e.g. SNPs or SNVs), that have been associated with a disease or some other binary phenotypic trait. Following Purcell et al 2009, Schizophrenia Working Group 2014, define the allelic PRS in the context of a dichotomous trait for an individual as

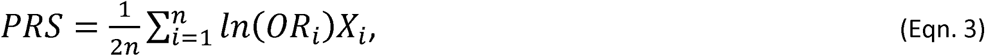

where *n* is the number of SNPs (sites) included in the PRS, *OR_i_* is the allelic case/control odds ratio for the *i^th^* SNP, and *X_i_* is the number of disease risk alleles that the individual carriers at the *i^th^* SNP (0, 1, or 2). In the definition above, *OR_i_* = *f_cs_*(*f_ct_*)^-l^(1- *f_ct_*)(1- *f_cs_*)^-l^; where *f_cs_* is the frequency of the disease risk allele at the *i^th^* SNP within cases and *f_ct_* is the frequency of that allele within the controls. The factor of 2 in the denominator accounts for diploidy and *max(X*_i_) = 2 at each site. There are other proposed heuristic metrics for PRS, such those with *OR* in rather than *ln(OR)* in Eq 3, but our study focuses exclusively on this definition, both because it is the most widely-used in the literature, and because this definition is congruent with the coefficients of a logistic predictor model of disease from genotypes (e.g. Wray et al 2010, Dudbridge 2013).

### Monte Carlo Simulation Construction

To generate a large set of genetic markers with variable disease-susceptibility effects under models described above, we constructed a Monte Carlo simulation of disease risk in multilocus genotype samples. We simulated a scenario where risk-associated SNPs had been identified by GWAS, so that the majority of simulated sites carry risk alleles while a much smaller fraction are null sites (false positives in GWAS) which do not contribute to disease risk. We do not simulate the scenario of exploratory random assays of (potentially) millions of SNPs where the great majority would have null effects and only a very small fraction contribute to disease risk.

Our experimental design for assessing the performance of classifiers in relation to inheritance model and penetrance γ values involved simulating n = 2000 disease and 2000 control multilocus genomes in both the training and the test sets, so that the set of simulated genomes was balanced with respect to both disease/control and training/test set sample size (i.e. with a total of 8000 genotypes for every simulation). Each genome in the simulation has L = 500 risk + 50 null polymorphic sites. As noted, the total number of SNPs was selected to be consistent (within an order of magnitude) with the number of sites identified in association with many common diseases by GWAS (e.g. Fritsche et al 2016), while the fraction of null sites was (conservatively) selected to be somewhat higher than the standard false discovery rate (FDR) of 0.05.

The genotype at each site of a simulated genome is assigned by sampling from the conditional probabilities of genotype given disease or control phenotype in Eqs 1-2, with the underlying assumption of Hardy-Weinberg equilibrium frequencies for the diploid genotypes at each locus together with a beta distribution parameterization of allele frequencies. Genotypes are assigned at each site independently of the alleles at all other loci, in keeping with the assumption of linkage equilibrium among all simulated loci. We assume a baseline “wild type” disease rate *f*_11_ = 0.01, so that the probability of developing the disease in individuals with no risk alleles is 0.01. Deviations from this baseline due to the effects of risk alleles are determined by the penetrance parameter γ= 1 + δ, defined above in the disease model, where the penetrance perturbation value o is a uniform random variable on 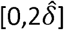, with 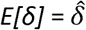. We simulate genotypic risk over 16 values of 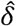 ranging from 0 to 0.0075 in intervals of 0.005, with 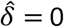 corresponding to the baseline risk of 0.01 for all genotypes and replicates while, for comparison, e.g. 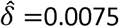 generates disease risk as a uniform random variable in the range 0.01 to 0.025 (or, on average an expected 75% increase in disease risk in comparison to the baseline). The upper bound of 0.0075 was selected because in preliminary simulations, values of 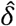 larger than this were associated with unrealistically high predictive accuracies (approaching 1.0) and thus provided little information.

The contributions of the perturbation parameter (measuring penetrance) to disease risk under additive, recessive, and dominant inheritance were parameterized as described in the previous section. In addition to modeling all 500 risk alleles as either completely additive, recessive, or dominant in their effects on disease phenotype, we also consider scenarios of mixed effects, i.e. 400 sites with additive effects + 100 sites with recessive alleles as well as the scenario 400 additive + 100 dominant effect risk allele sites. These mixed models were introduced to determine whether a subset of sites with individually strong or weak effects would primarily determine the performance of classifier algorithms at predicting disease phenotype.

### Prediction Modeling

From each iteration of the Monte Carlo simulation, the generated multilocus genotypes composed of disease risk and null SNPs were used as features (predictor variables) for the development of machine learning models to predict disease status. The models were parameterized using the training set with equal numbers of case and control genomes as described above. One set of analyses is carried out on a subset of sites filtered using LASSO (Tibshirani 1996) for feature selection, and a second set of analyses was run without LASSO. This approach allows a direct comparison of the performance of the classification machine-learning algorithms on LASSO-filtered data sets to those performed without prior feature selection. LASSO was implemented using the R 4.3.0 glmnet package (Friedman et al 2010, Simon et al 2011), selecting the penalty parameter λ from the range [0,0.05] in increments of 0.01. The feature selection had the additional constraint of requiring least 10% of the sites being retained for analysis – those that failed to return at least this fraction of sites were excluded from analysis.

All eight predictive combined scenarios (2 in the case of analyses with and without feature selection x 4 predictive models) were applied to data generated under a series of additive, dominant and recessive disease models (as well as the mixed inheritance models) to assess their binary classification performance. The predictive models used were: i. *Logistic Regression* (*LR*, Hosmer and Lemeshow 1989), ii. *Random Forest* (*RF*, Ho 1995), iii. *Naïve Bayes* (*NB*, Domingos and Pazzani 1997) and iv. *Neural Network* (*NN*, James et al 2021, McCulloch and Pitts 1943) models to evaluate an array of different classifiers for the simulated data sets.

The RF and NB algorithms were implemented using the respective R packages randomForest (Liaw and Wiener 2002) and e1071 (Meyer et al 2024). We initially implemented neural network models using the R neuralnet library, but following a high failure rate in model convergence in preliminary analyses with < 10 replicates, we instead ran the h2o.deeplearning program (a feedforward multi-layer neural network, Candel et al 2022), by interfacing R with h2o.ai (2022). The specific NN model used was a multilayer feedforward NN. Also, due to inconsistency across replicates with the standard R glm function, the h2o.glm function was used for predictive modeling with logistic regression. Unless otherwise noted, the default parameterizations and settings of each classifier in R and h2o were used to analyze the simulated genomes.

### Evaluation

The simulated genotype – disease phenotype training and test set data were analyzed using LR, NB, RF, and NNs as described above. For each model, a tally of the number of true positives measured against the number of false positives is used to construct a Receiver Operating Characteristic (ROC) curve. The area under the curve (AUC) is a summary statistic of performance. The AUC can be interpreted as the probability that any two individuals, one with the disease and the other control, are correctly classified based on their genotypes. All AUC calculations were performed using the R pROC package (Robin et al 2011).

The relationship between AUC and PRS was evaluated by computing the mean PRS within the case and control groups and assessing how ΔPRS = PRS_case_ – PRS_control_ (with a mean computed over all cases and controls per replicate) scales against AUC for a LR predictor. This ΔPRS vs. AUC relationship was assessed and comparisons made across modes of inheritance with additive, recessive, and dominant effects (mixed effects were not considered for the PRS vs. AUC comparisons). The Pearson correlation between AUC and ΔPRS was calculated to determine the strength of association between these summary statistics.

As part of our analysis of the relationship between predictive accuracy and the difference between case and control PRS in our simulated data, we compared the AUC observed for ΔPRS values generated in the simulations to the analytical predictions of AUC from the mean and variance of PRS in Dudbridge (2013). Dudbridge derived AUC for logistic regression models under two scenarios of additive genetic risk effects. The first scenario assumes that there is a hidden, normally distributed “liability variable” determined by additive effects across multiple loci. When this liability variable exceeds a threshold value (determined by the disease prevalence in the population and the assumption of normality of the liability variable), a disease phenotype results. Interpreting PRS as the coefficients of the liability function in a linear model, Dubridge showed that the expected predictive accuracy given log odds PRS of the liability model is:

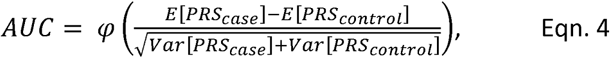

where *ψ* is the cumulative density function of the standard normal distribution. For point of comparison with the simulations, our ΔPRS is a point estimator for the numerator term, the denominator is calculated from the observed variance in PRS across the 2000 case and control genomes.

Dudbridge also considered an alternative model where the PRS of an individual is the log risk of the disease for that genotype, rather than having PRS as an underlying liability variable with a threshold effect on disease risk. Assuming that the disease is rare in the population (i.e. background frequency << 1) and equal variances in PRS for both case and control samples, the log-risk model has an AUC estimator of:

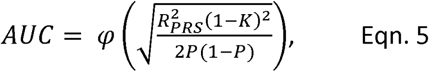

where 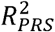 is the coefficient of determination of PRS on disease or control phenotype (computed from a simple linear regression of PRS in each replicate vs. binary disease phenotype using the *lm* function in R), *K* is the population prevalence of the disease (0.01 in our simulations), and *P* is the sample prevalence of the disease (0.5 in our model due to balanced sampling). In this study, we compare estimators of AUC in Eqs. 4–5 against the observed AUC under a logistic regression model for a range of PRS at different values of per-site genetic risk.

## RESULTS

There are several broad trends in our results concerning the association between penetrance values and AUC (Figures 1A-D, S1), and between AUC the heuristic PRS metric (Figure 2A-C). As expected, AUC is effectively 0.5 for 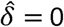 and monotonically increases with larger values of 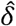 for all of the classifiers. Qualitatively, the general pattern of association between AUC and 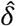 is one of approximately linear increase for values near zero before asymptotically approaching 1 for large penetrance for 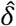 values, resembling a sigmoidal curve. The fastest rate of increase in AUC with respect to penetrance (i.e. highest AUC for small penetrance values) occurs with dominant inheritance (e.g. with dominant inheritance, AUC attains values ≥ 0.9 for 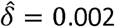 with all classifiers apart from NN (Fig 1B), whereas for additive (1A) and recessive effects (1C), this AUC value is not achieved for 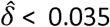 and 0.0035-0.004, respectively, depending on the classifier).

**Figure 1.**
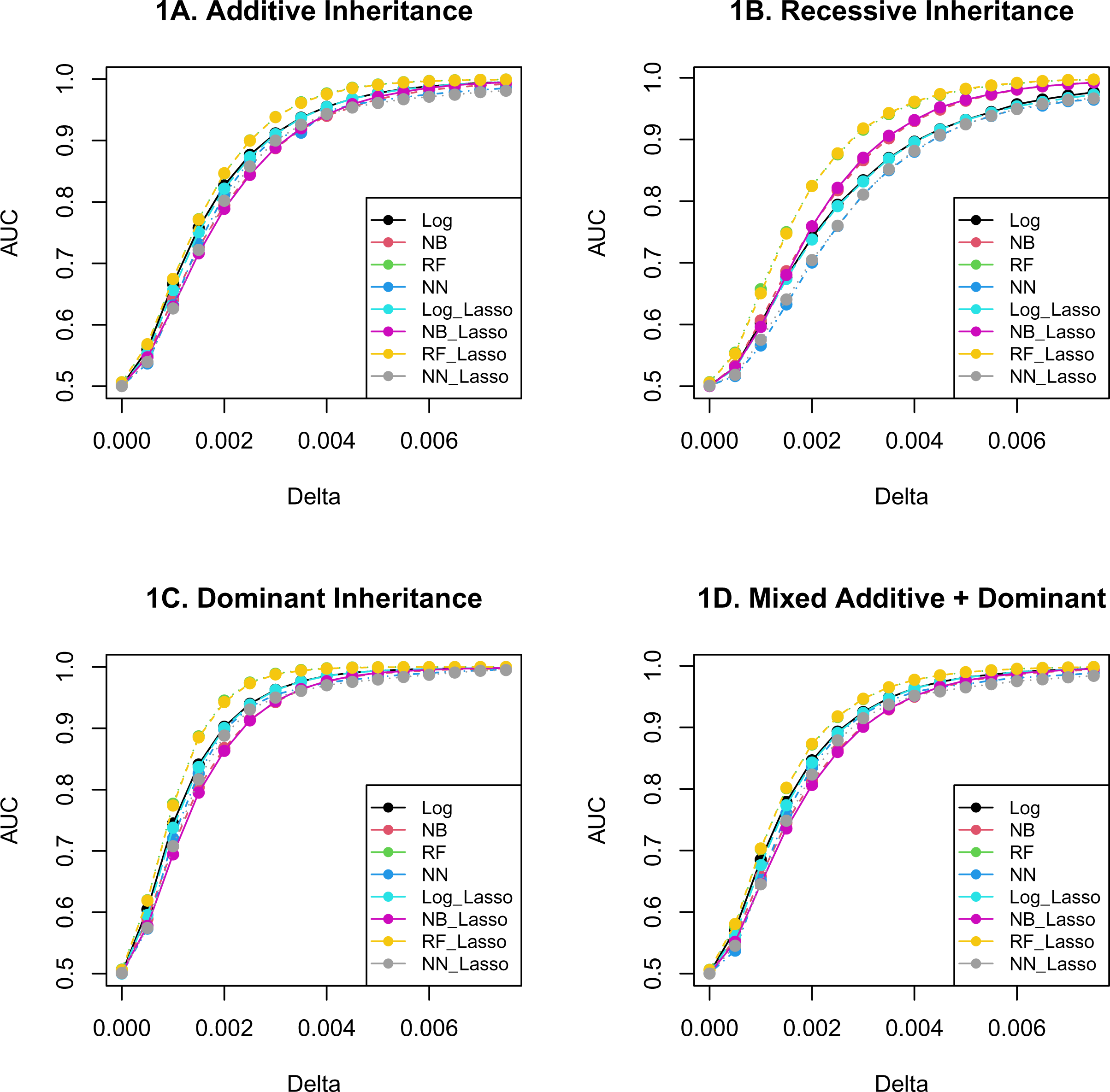
AUC vs. penetrance 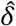 for various models of genetic risk inheritance across several predictive models (logistic regression, naïve Bayes, random forests, and neural networks), contrasting results with and without LASSO-based feature selection. Note that RF models give the highest predictive accuracy while NN give the lowest for most values, as well as the fact that feature selection has little impact on AUC. The AUC scores shown are the mean values over 100 replicates. A. Additive model of disease risk inheritance. B. Recessive model of disease risk inheritance C. Dominant disease risk inheritance D. Mixed additive-dominant model of disease risk inheritance (400 additive, 100 dominant effect sites)

**Figure 2.**
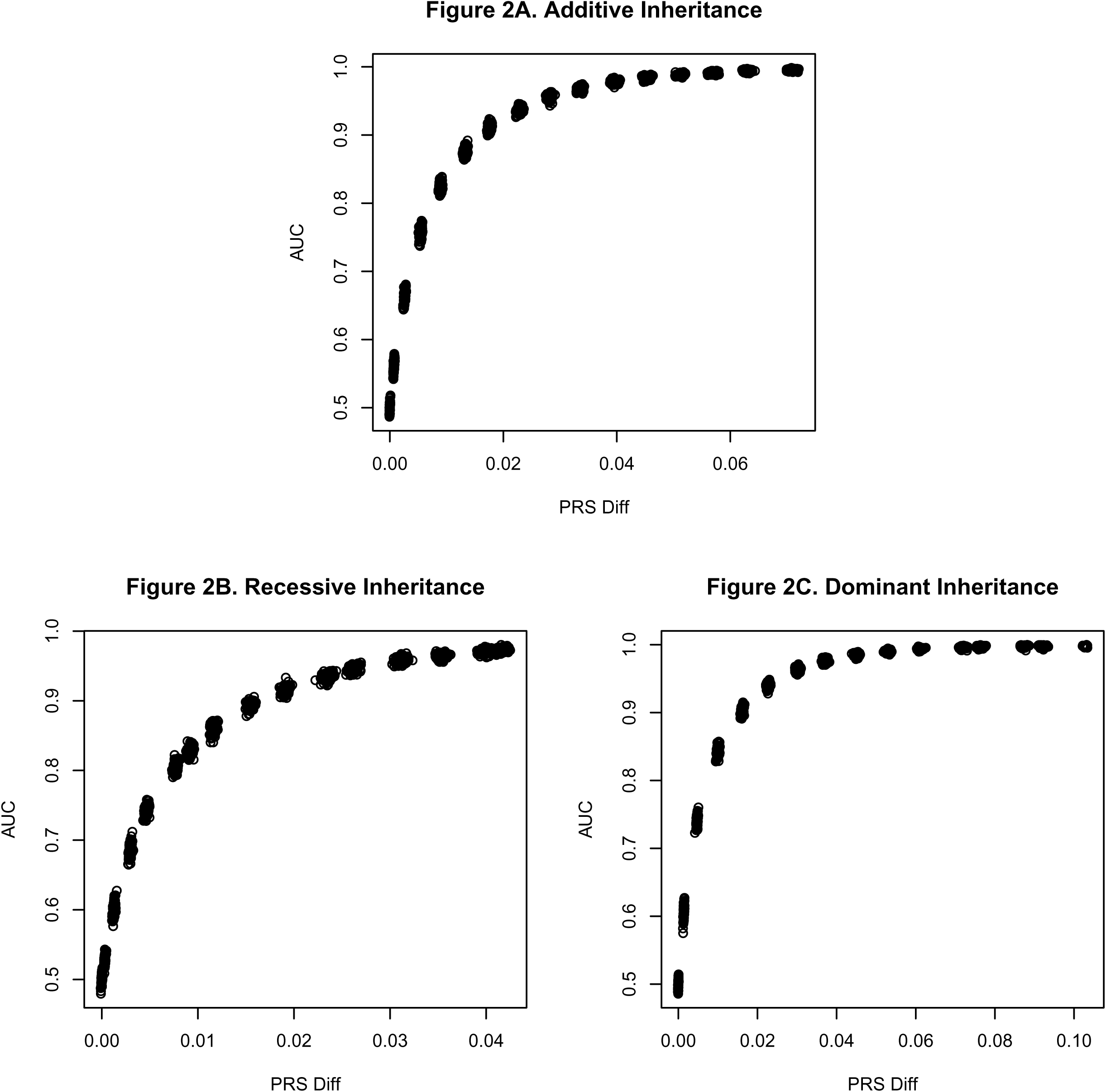
Plots of AUC vs. ΔPRS (the mean difference Polygenetic Risk Score between cases and control individuals) using logistic regression as a predictive model for A. additive, B. recessive, and C. dominant models of disease risk inheritance, with all 100 replicates per value included in the plots. The Pearson correlations between AUC and PRS are, for the three respective models of inheritance, 0.884 (additive), 0.924 (recessive), and 0.815 (dominant), with p << 1e-6 for all scenarios.

For all modes of inheritance and over most of the range of penetrance 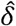 values, RF classifiers out-perform all other algorithms in correctly predicting disease status from genotype at intermediate 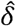 values. The greater predictive accuracy of RF in comparison to the other classifiers is especially pronounced with recessive effects of risk alleles, whereas for dominant effects the divergence of AUC among predictors is comparatively small. In contrast, the NN almost invariably had lower predictive accuracy when compared to the other predictive models – again with the greatest differences in performance in comparison to other classifiers observed with recessive inheritance. LR and NB are intermediate in performance between RF and NN. LR and NB give quite similar AUC values for additive and dominant inheritance, but in the case of recessive inheritance, they start to diverge for larger values of 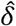, with LR consistently outperforming NB.

There is little improvement in classifier performance when including LASSO feature selection. The corresponding AUC values are either nearly identical, i.e. in Figure 1, and most of the AUC averages with or without feature selection are almost superimposed. Indeed, in many instances the mean AUC is slightly larger in the absence of LASSO (e.g. with additive inheritance, the mean difference in AUC for LR with and without AUC is 0.002, although this difference is not statistically significant, and, in contrast, the difference in mean AUC for NN under recessive inheritance with vs. without AUC is −0.002, which is also not statistically significant). These results presumably reflect the fact that only 10% of sites in the model are null, so that feature selection may remove risk sites as false negatives along with the true negative nulls for models with weak risk effects, while for larger penetrance values, the classifiers are unlikely to fail in capturing effects from risk sites with or without feature selection.

The AUC values for mixed-effect models, are, as expected, intermediate in comparison to pure additive and pure recessive or dominant, but more closely resemble the additive models. For example, if we compare the AUC for the RF classifier in the mixed additive + dominant model (Fig. 1D), we can see that the AUC attains a value > 0.9 at 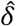 values between 0.002 and 0.0025, which is larger than the 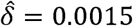 for the case where all risk alleles have dominant effects (Fig. 1D) but slightly lower than the 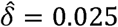 for the pure additive model. A similar intermediate pattern, though in the reverse direction, is observed for a mixed additive + recessive model (SI1). The AUC for mixed models more closely resembles the results for the pure additive effects than either the pure dominant or recessive effects, suggesting that the majority of sites in our model (400 out of 500 risk sites) with additive effects largely drive the predictive models and that the small subset of sites with non-additive inheritance effects do not appear to be the principal determinants of model performance, even when the non-additive minority have dominant effects.

The plots of AUC vs. ΔPRS for the same 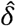 are consistent with PRS being a strong heuristic predictor of disease risk and of classifier performance for our genetic models. Specifically, there is a significant correlation between ΔPRS and the AUC classifier performance metric for all models of inheritance, reflecting the close correspondence between ΔPRS and AUC for each increment in penetrance value (Figure 2A-C for additive, recessive, and dominant inheritance – the dense clusters of points at regular intervals of the plot are the 100 replicates per penetrance increment).

As predicted, ΔPRS increases monotonically with 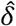 as does AUC, thus the correlations between ΔPRS and AUC are statistically significant despite the non-linearity and asymptotic convergence to 1.0. Specifically, the correlations range from > 0.8 for additive and recessive effects to > 0.76 for dominant inheritance, with p << 1e-6 for the correlation coefficients in all models. The faster rate of convergence to 1.0 for dominant inheritance reduces the approximately linear component of the AUC vs. ΔPRS relationship and accounts for the somewhat smaller but still significant correlation coefficient. Note the larger values of ΔPRS for the same set of 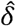 in the dominant inheritance model, i.e. a maximum < 100 in the recessive case vs. 180 in the dominant effects case, indicating a greater divergence in disease vs. control odds ratios in association with dominant effect risk alleles.

The relationship between mean AUC and mean ΔPRS (Figure 3) shows a pattern consistent with that of a normal cdf as derived in Dudbridge (2013) for an additive model of disease risk inheritance, at least within certain ranges of ΔPRS. Figure 3 contrasts the observed relationship between predictive accuracy and case-control difference in PRS to the to the values of AUC predicted from the mean and variances of PRS in Eqs 4-5. As expected, the variances in PRS among case and control are approximately normally distributed (p >> 0.05 for Kolmogorov-Smirnov test) and are nearly identical for small 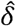, e.g. case and control variance in PRS = 1.81e-6 and 1.82e-6 at 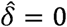. For large delta, the values are slightly divergent, e.g. for 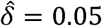, the respective case and control PRS variances are 1.37e-4, 1.19e-4.

**Figure 3.**
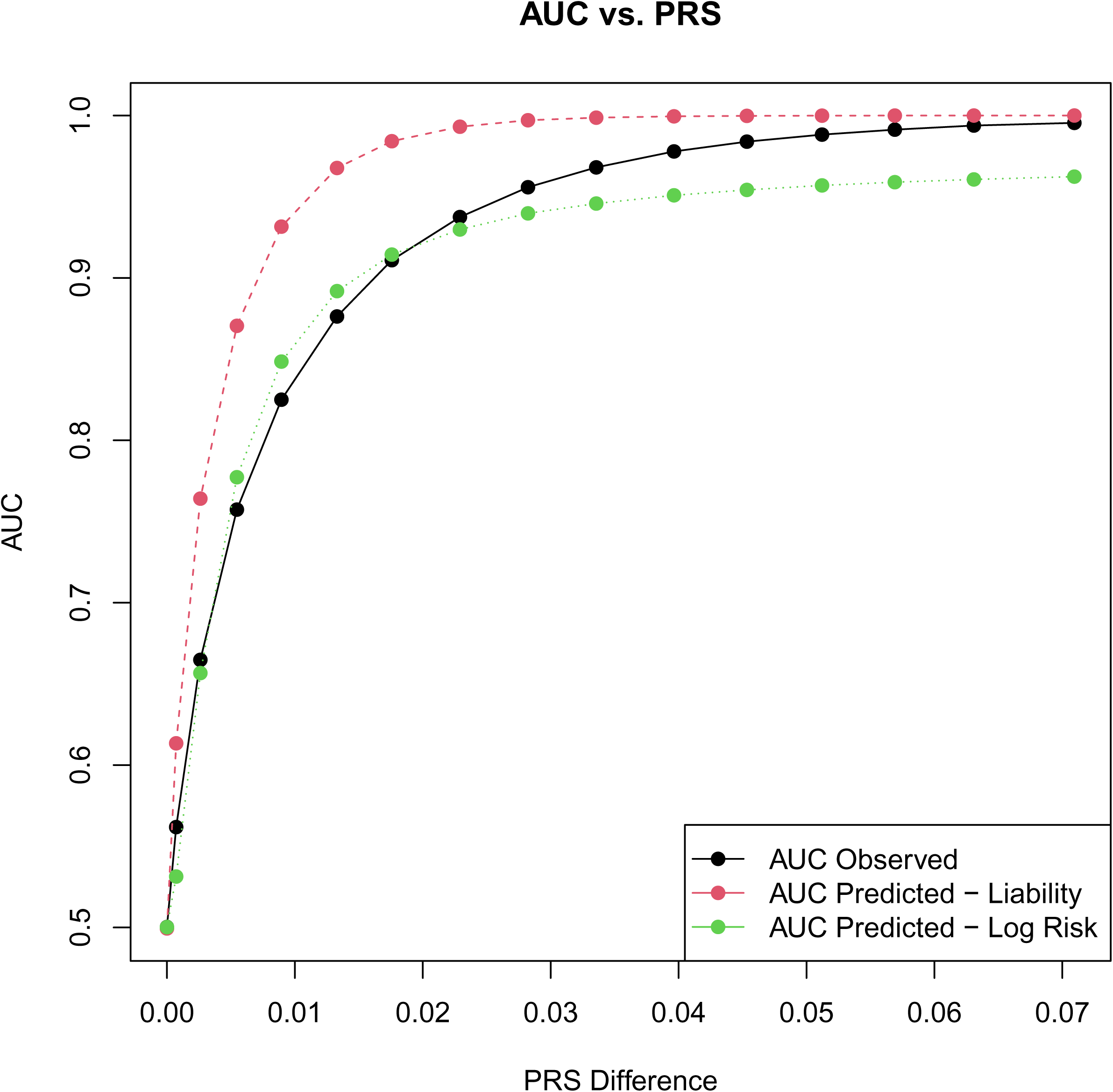
AUC vs ΔPRS under a logistic regression predictor and an additive model of disease risk. The observed AUC (black) are similar to the predicted AUC (green) under a log risk model for smaller values of ΔPRS. The log risk predicted values (computed from R2 between PRS and phenotype) fail to converge to AUC values near 1.0 for larger deviations in PRS. In contrast, AUC predicted from PRS under a liability threshold model (red) is consistently higher than the AUC observed in our model, except asymptotically for very large ΔPRS.

The AUC derived for a log-risk model for PRS (Eq. 5) provides a closer fit to the AUC vs. ΔPRS curve when the differences in PRS between case and control are small. However, for larger values of ΔPRS, the analytic approximation asymptotically tends to 0.96 rather than to 1.0 for the observed means and variances in PRS. Furthermore, even though the liability threshold model (Eq. 4) generates a comparatively poor fit for small (but non-zero) values of ΔPRS and 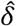, it does have the desired property of converging to 1.0 for the range of parameters considered.

Equations 4-5 were derived specifically for an additive model of disease risk, and thus would not precisely apply to scenarios with recessive or dominant effects. Nevertheless, as can be seen in the S2-A figures for dominant effects, the log-risk model still provides a relatively close approximation to the observed AUC when ΔPRS is small, while the liability threshold model AUC converges to the simulated values at larger ΔPRS values as in the additive case. Meanwhile, with recessive risk effects (S2-B), the log-risk model provides a better approximation for all ΔPRS values, because even at large ΔPRS the observed AUC converge to values < 1.0.

## SUMMARY AND DISCUSSION

Disease genetics modeling is an understudied approach for understanding the implications and limitations of GWAS analyses and their implications for disease diagnostics. Because there have been few systematic studies to determine which types of classifiers and feature selection algorithms performed well for different disease models, this study 1) provides a framework for conducting Monte Carlo simulations to better elucidate the performance of standard machine learning approaches to genetic-based prediction, 2) shows that, at least for this class of non-epistatic models of genetic architecture, random forests seemed to provide the highest predictive accuracy, while other classifier models such as logistic regression and naïve Bayes also effectively predicted disease phenotype from genotype under the various models of inheritance, and 3) provides evidence that PRS is a valuable predictive tool for quantifying disease risk from genetic data, as evidenced by the correlation between model AUC and the separation in average PRS between case and control genomes, supporting their use as a disease risk metric.

This study also found, unsurprisingly, that dominant effects of risk alleles have higher diagnostic utility signal compared to those with additive effects, which in turn give higher AUC for the same perturbation values than under a recessive model of inheritance. These results are also consistent with the greater ΔPRS between case and control seen with dominant inheritance, as well as with the fact that the highest divergence in classifier performance is seen with recessive inheritance, suggesting that while all classifiers can effectively identify the strong association with dominant effects, differences in classifier model accuracy start to become more apparent when the predictor effects are weaker and/or rarer. This runs counter to some heuristic arguments suggesting that sites with recessive effects can sometimes generate the highest genotype-based odds ratios between disease and control (e.g. aa for disease vs. AA or Aa control) on the grounds that genotypes associated with disease would be particularly rare and distinctive, as opposed to cases where risk alleles in the heterozygous state could be found in both case or control.

The fact that RF had the highest predictive accuracy while NN had the lowest under a wide range of models and parameter values is particularly interesting in view of the fact that both NN and RF (at least implicitly) create models that assign a higher weight to some sites than to others, in contrast to LR and NB that treat each site independently and thus of equal weight. Consequently, one might expect NN and RF to have performances more similar to one another than to LR and NB, either improving upon the independent models by correctly assigning higher weight to more informative predictive sites or having reduced performance by assigning high weights to false positives (e.g. null sites or low risk sites) generated by stochastic correlations. In fact, the divergences in AUC suggest that RF correctly creates decision trees with informative sites at the top of the hierarchy, whereas the NN seem to be assigning higher weights to uninformative sites at a higher rate, at least for simple models without linkage disequilibrium or epistatic interactions.

The observation that feature selection via LASSO has a negligible contribution to predictive model performance, as seen from the fact that mean AUC with or without LASSO for the same classifier are nearly identical, was somewhat surprising. This probably largely reflects the fact that we model a scenario where null sites are small fraction (< 10%) of the total number of SNPs, so that the number of false positives such sites can contribute is negligible compared to the associations with informative sites. In a scenario where the majority of sites were null - i.e. one where the set of loci was part of an exploratory study rather than identified by a prior GWAS – feature selection would probably play a much more important role in efficiently generating accurate predictive models.

Additionally, our simulations provide further support to the use of PRS as an indicator of disease risk, due to the strong association between ΔPRS and AUC both with additive and recessive/dominant effects. Apart from the heuristic and intuitive appeal of PRS, its association with AUC in the case of additive genetic architectures under a LR predictive model was shown analytically in Dudbridge (2013).

The good fit of AUC in Eqn. 5 to our simulated values in the lower range of ΔPRS implies that with small 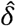 and small divergence in PRS between disease and control individuals, a log risk interpretation of PRS is a good model for disease prediction. In contrast, the divergence between observed AUC and that predicted from the log risk model for larger values of 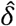 and ΔPRS suggest that some of the log risk model assumptions start to break down, as seen in the model’s failure to asymptotically converge to AUC = 1.0. The variances in PRS in the case and control samples diverge for the larger ΔPRS, violating the assumptions in the derivation of Eq. 5 (furthermore, an inflated error rate increases the sample variances of PRS and reduces AUC). The closer fit of Eq. 4 to the simulated AUC for high ΔPRS implies that for larger risk effects, a liability threshold model may be a more effective approximation for disease risk. This distinction also seen from our result where the log risk model in Eq. 5 more closely approximates AUC for larger values of ΔPRS when the risk effects are recessive rather than dominant (or additive), i.e. S2.

A possible reason for why the log risk model is a better predictor of AUC for small 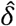 while the liability model more closely approximates the simulated data for large 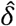 is that for larger effects, the variance in genetic risk in the general population increases. This results in a sharper partitioning between disease and control phenotypes with respect to their genotypes and their respective numbers of risk alleles, thus approximating a threshold liability model. In contrast, when 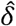 and the variance in genetic risk are small, the genetic distances between disease and control individuals tend to be small as well, resulting in considerable overlap in sets of risk alleles between disease and control genotypes. Under such a scenario, the liability model with its sharp threshold effect is not a good approximation to AUC due to the high overlap in genotypes between case and control, while a log risk model would be more effective at predicting this less discretized disease incidence.

We remark that while we only fit AUC vs. PRS models for LR because of the analytical relationship between PRS and logistic regression coefficients derived by Dudbridge, the shape of the curves for the associations between AUC and 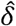 are consistent across predictive models, and it can be surmised that the correlations between PRS and AUC would be significant and a good fit under other classes of predictive models.

While our results are encouraging in demonstrating the ability of commonly-used classifier algorithms to predict disease phenotype from risk allele genotype, there are several limitations to this initial study of predictive model performance evaluation across disease models, suggesting future directions with more complex and biologically realistic scenarios. First, our simulations generate each SNP in linkage equilibrium with respect to all other sites. This is a reasonable assumption for modeling unique disease susceptibility loci across the genome, due to the fact that many GWAS studies identify sets of SNPs where the overwhelming majority have LD near 0, and because SNP panels generated from GWAS studies for PRS-based disease prediction typically only use a small fraction of sites with leading effects, ignoring second-order effects due to their associations with other sites.

However, incorporation of linkage disequilibrium patterns and other correlational effects, such as epistatic interactions and LD among loci contributing to disease risk, would further generalize the results, as it is known that significant LD occurs among many 10-100 kb regions of human chromosome, with D much higher than predicted under neutral evolution (e.g. Collins et al 1999, Pritchard and Przeworki 2001). The assumption of linkage equilibrium is what enabled logistic regression and the naïve Bayes classifiers to outperform a neural network model, which, unlike NB and LR does not explicitly assume conditional independence between features. In contrast, if the underlying assumptions of statistical independence are violated by linkage disequilibria or by genetic architectures with strong epistatic components, one may expect that neural networks would outperform classifiers such as LR or NB because of their ability to incorporate joint associations among sites as part of their predictive models.

Finally, we will note that although this study produced useful preliminary results for understanding the performance of different classifiers using disease genetic data, the size of the simulation can be expanded in the number of SNPs which would also improve the modeling of genomes, as well as in the sample size of individuals. While our aim was to initiate the investigation into the use of disease genetic models to identify those predictive approaches that produce high AUCs from genetic features, additional work in this area expanding the disease models, size of the study, and methods to evaluate discrimination performance will further advance this line of inquiry.

## SUPPLEMENTARY INFORMATION

**S1.** AUC vs. penetrance o for a mixed inheritance model (400 additive, 100 recessive effect sites)

S2. AUC vs. PRS for dominant (A) and recessive (B) models of inheritance (in black), compared to predicted values of AUC from PRS values under an additive model of inheritance. As in Figure 3, the green line shows predicted values with a log odds risk model, the red line predicted AUC with a liability threshold model.

## DATA AVAILABILITY

The R code for the Monte Carlo simulation is publicly available at https://github.com/mshpak76/Genetic_Disease_Simulation/

## STATEMENTS AND DECLARATIONS

### Author Contributions

EP, MS, and SM wrote the simulation and classifier code, MS analyzed the simulation output, SJS developed the research project, SJS and MS wrote the paper, SM and MM provided expertise on the applicability of the machine learning-based classifiers used in the study.

### Funding

Start-up funds from the Laboratory of Genetics, School of Medicine and Public Health, Office of the Vice Chancellor for Research and Graduate Education, and the Center for Human Genomics and Precision Medicine at the University of Wisconsin at Madison were used to support this study.

### Conflict of Interest

Eric Parfitt is employed by Wolfram Research, Inc. The remaining authors declare that the research was conducted in the absence of any commercial or financial relationships that could be construed as a potential conflict of interest.

## REFERENCES

Alzoubi H, et al. (2023) Deep learning framework for complex disease risk prediction using genomic variations. Sensors 23:4439.

Baker M, et al. (2019) Maximizing the benefit of life-saving treatments for Pompe disease, spinal muscular atrophy, and Duchenne muscular dystrophy through newborn screening. JAMA Neurol. 76:978–983.

Beck T, Shorter T, Brookes AJ (2020) GWAS Central: a comprehensive resource for the discovery and comparison of genotype and phenotype data from genome-wide association studies. Nucleic Acids Res. 48(D1):D933–D940.

Bracher-Smith M, et al. (2021) Machine learning for genetic prediction of psychiatric disorders: a systematic review. Mol Psychiatry 26:70–79.

Buniello A, MacArthur JAL, Cerezo M, et al. (2019) The NHGRI-EBI GWAS Catalog of published genome-wide association studies, targeted arrays and summary statistics. Nucleic Acids Res, Vol. 47 (Database issue): D1005–D1012.

Burt R and Neklason DW. (2005) Genetic testing for inherited colon cancer. Gastroenterology 128:1696–1716.

Candel, A., Parmar, V., LeDell, E., and Arora, A. (Nov 2024). Deep Learning with H2O. http://h2o.ai/resources.

Choi SW, Mak TS, O’Reilly PF (2020) Tutorial: a guide to performing polygenic risk score analyses. Nat Protoc/ 15: 2759–2772.

Collins A, Lonjou C, Morton NE (1999). Genetic epidemiology of single-nucleotide polymorphisms. Proc Natl Acad Sci USA, 96: 15173–15177.

De La Vega FM, Bustamante CD 2018. Polygenic risk scores: a biased prediction? Genome Med. 10:100.

Domingos P and Pazzani M. (1997) On the optimality of the simple Bayesian classifier under zero-one loss. Machine Learning 29:103–137.

Dudbridge F. (2013) Power and predictive accuracy of polygenic risk scores. PLoS Genet. 9:e1003348.

El-Husseini ZW, Gosens R, Dekker F, Koppelman GH (2020) The genetics of asthma and the promise of genomics-guided drug target discovery. Lancet Respir Med. 8:1045–1056.

Ewens WJ (1979) Mathematical Population Genetics. Springer-Verlag. Berlin Heidelberg Germany.

Farnaes L, et al. (2018) Rapid whole-genome sequencing decreases infant morbidity and cost of hospitalization. NPJ Genom Med. 3:10.

Friedman JH, Hastie T, Tibshirani R (2010) Regularization paths for general linear models via coordinate descent. Jour Stat Software 33:1–22.

Fritsche LG, et al. (2016) A large genome-wide association study of age-related macular degeneration highlights contributions of rare and common alleles. Nat Genet. 48:134–143.

Gallagher MD, Chen-Plotkin AS (2018) The post-GWAS era: from association to function. Am J Hum Genet. 102:717–730.

Ghafouri-Fard S, et al. (2020) Application of artificial neural network for prediction of risk of multiple sclerosis based on single nucleotide polymorphism genotypes. J Mol Neurosci. 70:1081–1987.

Ho D, et al. (2019) Machine learning SNP based prediction for precision medicine. Front Genet. 10:267.

Ho TK (1995) Random decision forests. Proceedings of 3rd International Conference on Document Analysis and Recognition, 1: 278–282.

Hosmer DW and Lemeshow S. (1989) Applied Logistic Regression. New York, Wiley.

H2O.ai. (2022) h2o: R Interface for H2O. R package version 3.42.0.2. https://github.com/h2oai/h2o-3.

James G, Witten D, Hastie T, Tibshirani R. (2021) An Introduction to Statistical Learning, 2nd Ed. Springer Nature.

Janssens AC, Aulchenko YS, Elefante S et al 2006. Predictive testing for complex diseases using multiple genes: fact or fiction? Genet Med. 8: 395–400.

Janssens AC, et al. (2007) The impact of genotype frequencies on the clinical validity of genomic profiling for predicting common chronic diseases. Genet Med. 9:528–535.

Jansenns AC (2019). Validity of polygenic risk scores: are we measuring what we think we are? Hum Mol Genet. 28:R143–50.

Liaw A, Wiener M (2002). Classification and Regression by randomForest. R News, 2(3), 18–22.

Levey DF, Stein MB, Wendt FR, Pathak GA, et al. (2021) Bi-ancestral depression GWAS in the Million Veteran Program and meta-analysis in >1.2 million individuals highlight new therapeutic directions. Nat Neurosci. 24: 954–963.

Lewis CM, Vassos E (2020). Polygenic risk scores: from research tools to clinical instruments. Genome Med. 12:44.

Liaw A, Wiener M (2002). Classification and regression by randomForest. The R Journal 2:18–22.

Maadooliat M, et al. (2016). The decay of disease association with declining linkage disequilibrium: a fine mapping theorem. Front. Genet. 7: 217

Maurano MT, Humert R, Rynes E, et al (2012) Systematic localization of common disease-associated variation in regulatory DNA. Science 337:1190–1195.

McCulloch W and Pitts W. (1943) A logical calculus of ideas immanent in nervous activity. Bull Math Biophys. 5:115–133.

McInnes G, Yee SW, Pershad Y, Altman RB (2021) Genomewide association studies in pharmacogenomics. Clin Pharmacol Ther. 110:637–648.

Meyer D, Dimitriadou E, Hornik K, Weingessel A, Leisch F (2024). e1071: Misc Functions of the Department of Statistics, Probability Theory Group (Formerly: E1071), TU Wien. R package version 1.7-16.

Moonesinghe R, et al. (2011) Discriminative accuracy of genomic profiling comparing multiplicative and additive risk models. Eur J Hum Genet. 19:180–185.

Nielsen DM, Ehm MG, Weir BS (1998) Detecting marker-disease association by testing for Hardy-Weinberg disequilibrium at a marker locus. Am J Hum Genet. 63:1531–1540.

Owen KR, et al. (2010) Assessment of high-sensitivity C-reactive protein levels as diagnostic discriminatory of maturity-onset diabetes of the young due to HNF1A mutations. Diabetes Care 33:1919–1924.

Patrick MT, et al. (2018) Genetic signature to provide robust risk assessment of psoriatic arthritis development in psoriasis patients. Nat. Commun. 8:4178.

Polygenic Risk Score Task Force of the International Common Disease Alliance. (2021) Responsible use of polygenic risk scores in the clinic: potential benefits, risks and gaps. Nat Med. 27:1876–1884.

Pritchard JK, Przeworki M (2001). Linkage disequilibrium in humans: models and data. Am J Hum Genet. 69:1–14.

Purcell SM, et al. (2009) Common polygenic variation contributes to risk of schizophrenia and bipolar disorder. Nature 460:748–752.

Reay WR, Cairns MJ (2021) Advancing the use of genome-wide association studies for drug repurposing. Nat Rev Genet 22:658–671.

Relling MV, et al. (2019) The Clinical Pharmacogenetics Implementation Consortium: 10 years later. Clin Pharmacol Ther. 107:171–175.

Robin X, Turck N, Hainard A, et al (2011). “pROC: an open-source package for R and S+ to analyze and compare ROC curves.” BMC Bioinformatics, 12, 77.

Roden DM, et al. (2018) Benefit of preemptive pharmacogenetic information on clinical outcome. Clin Pharmacol Ther. 103:787–794.

Samadder NJ, et al. (2020) Comparison of universal genetic testing vs guideline-directed targeted testing for patients with hereditary cancer syndrome. JAMA Oncol. 7:230–237.

Schizophrenia Working Group of the Psychiatric Genomics Consortium. (2014) Biological insights from 108 schizophrenia-associated genetic loci. Nature 511:421–427.

Schrodi SJ, Garcia VE, Rowland C, Jones HB. (2007) Pairwise linkage disequilibrium under disease models. Eur J Hum Genet. 15: 212–220.

Schrodi SJ, et al. (2014) Genetic-based prediction of disease trait: prediction is very difficult, especially about the future. Front Genet. 5:162.

Sham PC (1998) *Statistics in Human Genetics*. New York, Wiley.

Simon N, Friedman JH, Hastie T, Tibshirani R (2011). Regularizaton paths for Cox’s proportional hazards model via coordinate descent. Jour Stat Software 39:1–13.

Telenti A, et al. (2018) Deep learning of genomic variation and regulatory network data. Hum Mol Genet. 27(R1):R63–R71.

Tibshirani R. (1996) Regression shrinkage and selection via the lasso. J Royal Stat Soc Ser B. 58:267–288.

Visscher PM, Wray NR, Zhang Q, Sklar P, McCarthy MI, Brown MA, Yang J (2017) 10 years of GWAS discovery: biology function, and translation. Am J Hum Genet. 101:5–22.

Watanabe K, Stringer S, Frei O, et al (2019) A global overview of pleiotropy and genetic architecture in complex traits. Nature Genet. 51: 1339–1348.

Wray NR, Yang J, Goddard ME, Visscher PM (2010). The genetic interpretation of area under the ROC curve in genomic profiling. PLoS Genetic. 6:e1000864.

Yang Q, Khoury ML, Botto L et al. (2003) Improving the prediction of complex disease by testing for multiple disease-susceptibility genes. Am J Hum Genet. 72:636–649.

