## Supplementary figures and images for "Evaluating Genetic-Based Disease Prediction Approaches Through Simulation"

### Suppl1

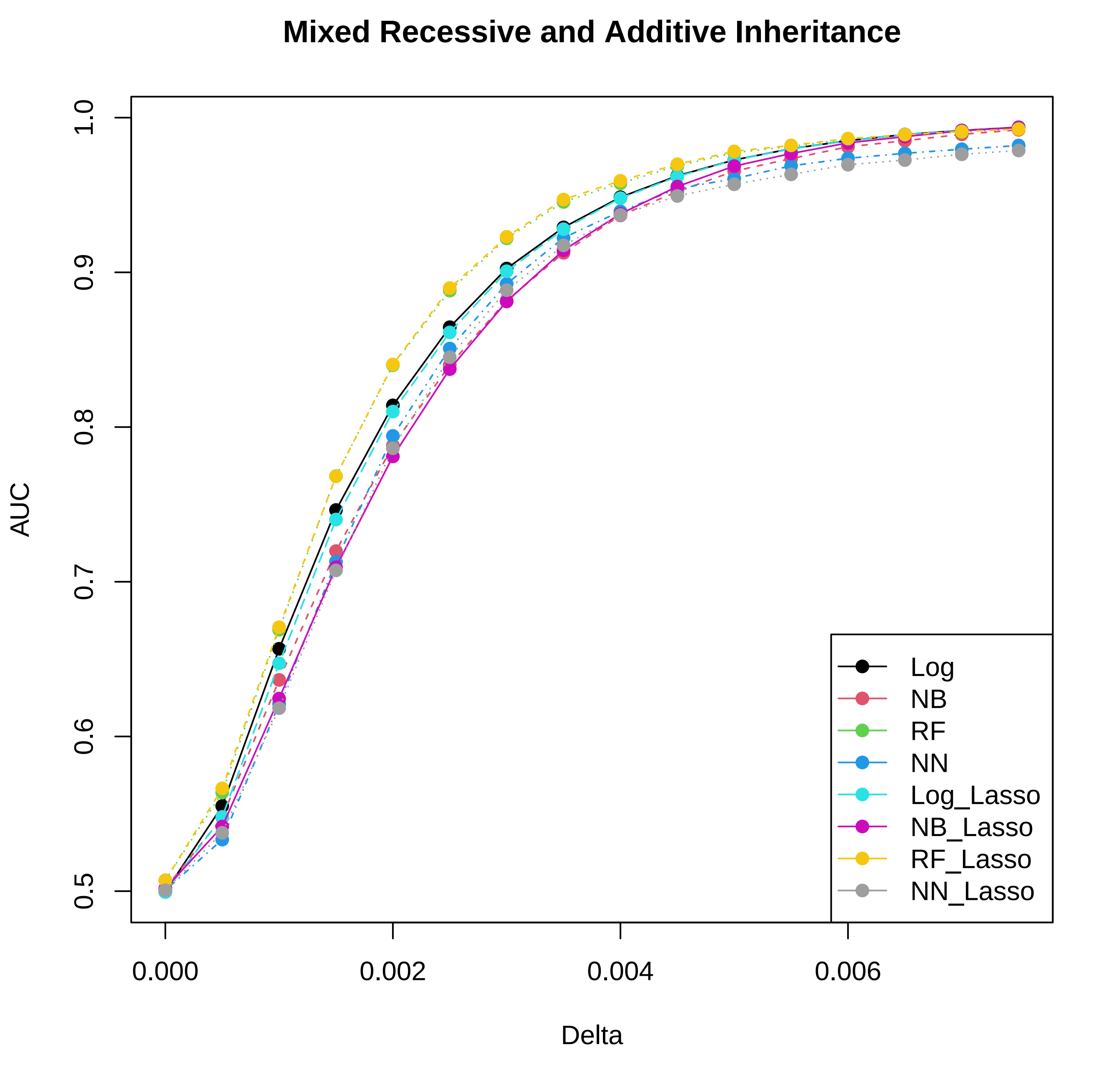

### Suppl2A

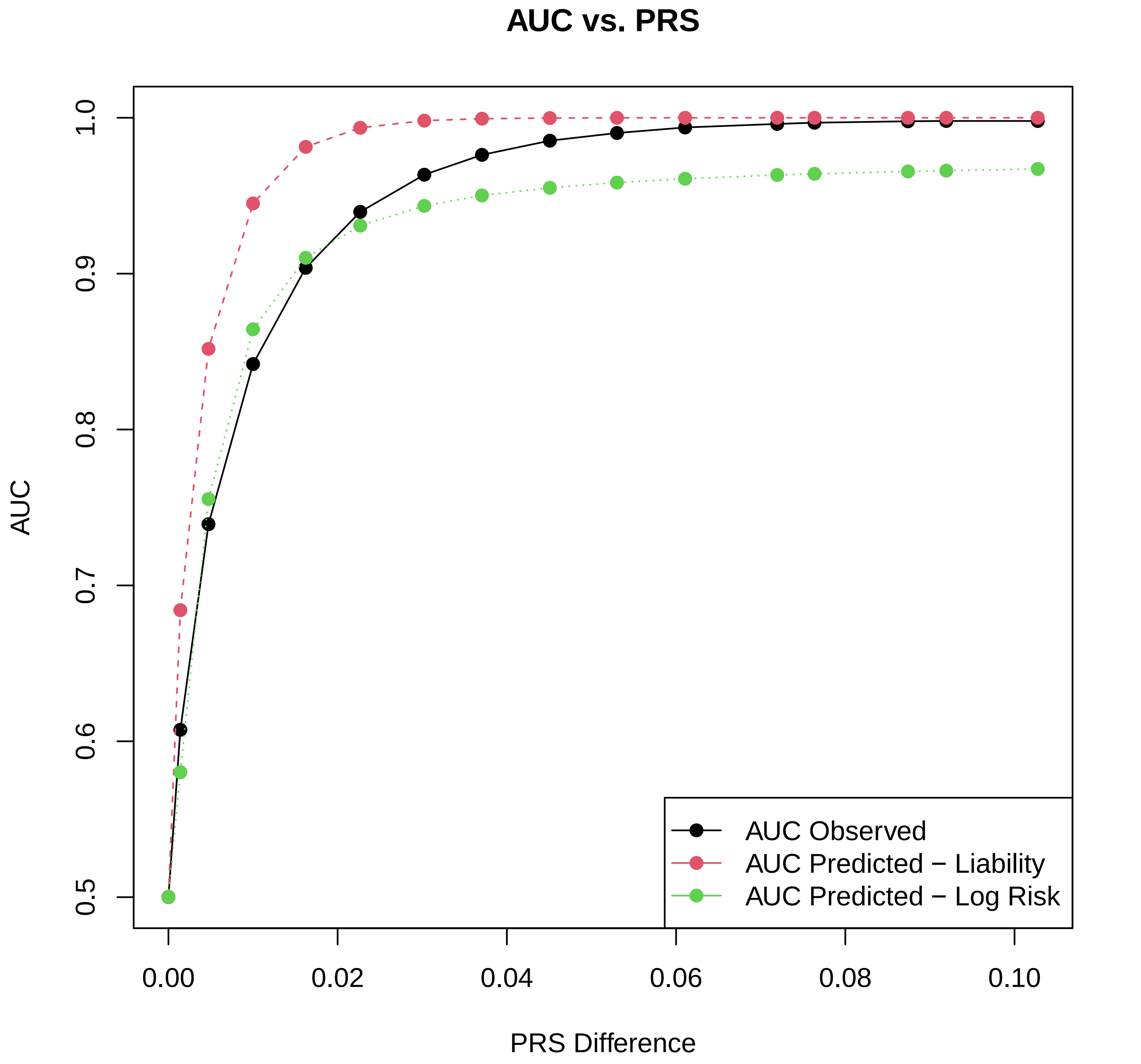

### Suppl2B

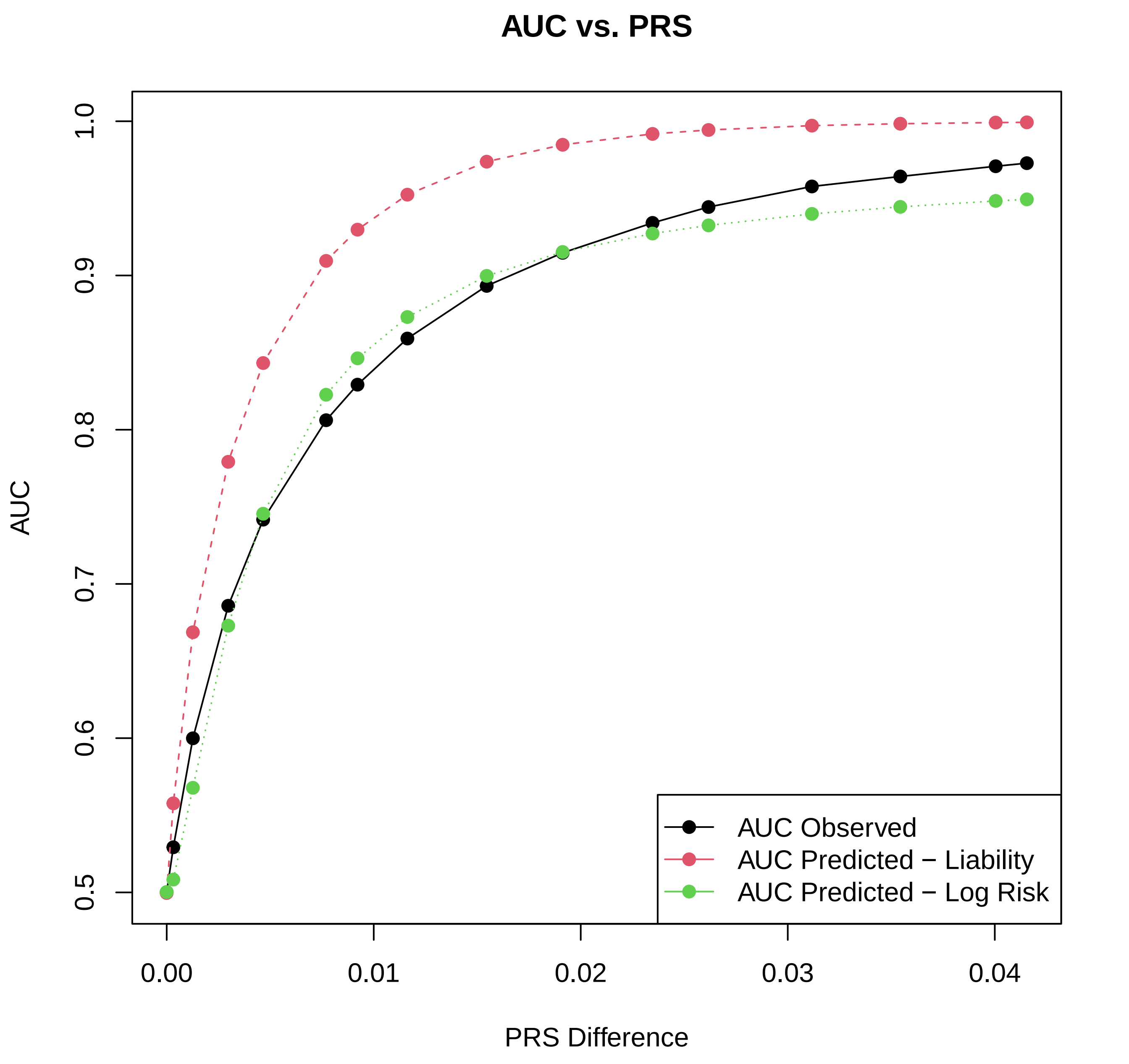
